# Hydroxychloroquine in patients mainly with mild to moderate COVID–19: an open–label, randomized, controlled trial

**DOI:** 10.1101/2020.04.10.20060558

**Authors:** Wei Tang, Zhujun Cao, Mingfeng Han, Zhengyan Wang, Junwen Chen, Wenjin Sun, Yaojie Wu, Wei Xiao, Shengyong Liu, Erzhen Chen, Wei Chen, Xiongbiao Wang, Jiuyong Yang, Jun Lin, Qingxia Zhao, Youqin Yan, Zhibin Xie, Dan Li, Yaofeng Yang, Leshan Liu, Jieming Qu, Guang Ning, Guochao Shi, Qing Xie

**Author notes:** Correspondence: Qing Xie, Department of Infectious Disease, Ruijin Hospital, Shanghai Jiao Tong University School of Medicine, 197 Ruijin 2nd Road, Shanghai, 200025, China, ORCID number: 0000-0002-2582-8803. Contributed equally and shared joint first authorship (Wei Tang, Zhujun Cao, Mingfeng Han, Zhengyan Wang). Contributed equally and shared joint corresponding authorship (Jieming Qu, Guang Ning, Guochao Shi, Qing Xie). Contributors: WT (Wei Tang), ZC (Zhujun Cao), MH (Mingfeng Han) and ZW (Zhengyan Wang) contributed equally to this paper and share joint first authorship. QX (Qing Xie), GS (Guochao Shi), GN (Guang Ning), and JQ (Jieming Qu) contributed equally to this paper and share joint corresponding authorship. QX and GS were co–chief investigators of this trial and act as guarantors of the study in its entirety. QX, GS, GN, and JQ led the development of the research question, study design, and obtaining the funding. WT, GS, QX, GN, JQ, and LL (Leshan Liu) coordinated the operational delivery of the study protocol to the coordinating centers. LL led the process of data cleaning and analysis and took responsibility for the results in this manuscript and future analysis. ZC led and produced the first draft of this manuscript. MH, ZW, JC (Junwen Chen), WS (Wenjin Sun), YW (Yaojie Wu), WX (Wei Xiao), SL (Shengyong Liu), EC (Erzhen Chen), WC (Wei Chen), XW (Xiongbiao Wang), JY (Jiuyong Yang), JL (Jun Lin), QZ (Qingxia Zhao), YY (Youqin Yan), ZX (Zhibin Xie), DL (Dan Li), and YY (Yaofeng Yang) represented the collaborating coordinating centers responsible for their centers’ participation in the trial. All authors provided critical review and final approval of the manuscript. The corresponding author attests that all listed authors meet authorship criteria and that no others meeting the criteria have been omitted.

## Abstract

**Objectives:** To assess the efficacy and safety of hydroxychloroquine (HCQ) plus standard–of–care (SOC) compared with SOC alone in adult patients with COVID–19.

**Design:** Multicenter, open–label, randomized controlled trial.

**Setting:** 16 government–designated COVID–19 treatment centers in China through 11 to 29 in February 2020.

**Participants:** 150 patients hospitalized with laboratory confirmed COVID–19 were included in the intention to treat analysis. 75 patients were assigned to HCQ plus SOC and 75 to SOC alone.

**Interventions:** HCQ was administrated with a loading dose of 1, 200 mg daily for three days followed by a maintained dose of 800 mg daily for the remaining days (total treatment duration: 2 or 3 weeks for mild/moderate or severe patients, respectively).

**Main outcome measures:** The primary outcome was whether participants had a negative conversion of SARS–CoV–2 by 28 days, and was analyzed according to the intention–to–treat principle. Adverse events were analyzed in the safety population in which HCQ recipients were participants who actually received at least one dose of HCQ and HCQ non–recipients were those actually managed with SOC alone.

**Results:** Among 150 patients, 148 were with mild to moderate disease and 2 were with severe disease. The mean days (± standard deviation, min to max) from symptoms onset to randomization was 16.6 (±10.5 days, 3 to 41 days). The negative conversion probability by 28 days in SOC plus HCQ group was 85.4% (95% confidence interval (CI) 73.8% to 93.8%), similar to that in the SOC group 81.3% (95%CI 71.2% to 89.6%). Between–group difference was 4.1% (95%CI –10.3% to 18.5%). In the safety population, adverse events were recorded in 7 (8.8%) HCQ non–recipients (N=80) and in 21 (30%) HCQ recipients (N=70). The most common adverse event in the HCQ recipients was diarrhea, reported in 7 (10%) patients. Two HCQ recipients reported serious adverse events.

**Conclusions:** The administration of HCQ did not result in a significantly higher negative conversion probability than SOC alone in patients mainly hospitalized with persistent mild to moderate COVID–19. Adverse events were higher in HCQ recipients than in HCQ non–recipients.

**Trial registration:** ChiCTR2000029868

**What is already known on this topic:**

— The pandemic of coronavirus disease 2019 (COVID–19) imposes substantial burdens on individuals, communities, health–care facilities, markets, governments, etc. globally.
— There is no specific treatment approved for COVID–19 or vaccine to prevent infection with the novel coronavirus.
— During the urgent pandemic, media headlines the utility of drugs without solid evidence but buries the side–effects of these drugs.

**What this study adds:**

— In this randomized clinical trial of patients mainly with persistent mild to moderate COVID–19, exposure to hydroxychloroquine led to a similar probability of virus elimination comparing to the current standard–of–care.
— Adverse events, mostly gastrointestinal related, were significantly increased in patients who received hydroxychloroquine.
— Overall, the results from our trial do not support the use of hydroxychloroquine in patients with persistent mild to moderate COVID–19.

**Print abstract:** *Study question:* To assess the efficacy and safety of hydroxychloroquine (HCQ) plus standard–of–care (SOC) compared with SOC alone in adult patients with COVID–19.

*Methods:* This is a multicenter, open–label, randomized controlled trial conducted in 16 government–designated COVID–19 treatment centers in China through 11 to 29 in February 2020. A total of 150 patients hospitalized with laboratory confirmed COVID–19 were included in the intention to treat analysis. Among them, 75 patients were assigned to HCQ plus SOC and 75 to SOC alone. HCQ was administrated with a loading dose of 1, 200 mg daily for three days followed by a maintained dose of 800 mg daily for the remaining days (total treatment duration: 2 or 3 weeks for mild/moderate or severe patients, respectively). The primary outcome was whether participants had a negative conversion of SARS–CoV–2 by 28 days, and was analyzed according to the intention to treat principle. Adverse events were analyzed in the safety population in which HCQ recipients were participants who actually received at least one dose of HCQ and HCQ non–recipients were those actually managed with SOC alone.

*Study answer and limitations:* Among 150 patients, 148 were with mild to moderate disease and 2 were with severe disease. The mean days (± standard deviation, min to max) from symptoms onset to randomization was 16.6 (±10.5 days, 3 to 41 days). The negative conversion probability by 28 days in SOC plus HCQ group was 85.4% (95% confidence interval (CI) 73.8% to 93.8%), similar to that in the SOC group 81.3% (95%CI 71.2% to 89.6%). Between–group difference was 4.1% (95%CI –10.3% to 18.5%). In the safety population, adverse events were recorded in 7 (8.8%) HCQ non–recipients (N=80) and in 21 (30%) HCQ recipients (N=70) with two serious adverse events. The most common adverse event in the HCQ recipients was diarrhea, reported in 7 (10%) patients. Two HCQ recipients reported serious adverse events.

*What this study adds:* Our trial does not support the use of hydroxychloroquine in patients with persistent mild to moderate COVID–19 due to limited effects on virus eliminating and significantly increased adverse events.

*Funding, competing interests, data sharing:* This work was supported by the Emergent Projects of National Science and Technology (2020YFC0844500), National Natural Science Foundation of China (81970020, 81770025), National Key Research and Development Program of China (2016YFC0901104), Shanghai Municipal Key Clinical Specialty (shslczdzk02202, shslczdzk01103), National Innovative Research Team of High–level Local Universities in Shanghai, Shanghai Key Discipline for Respiratory Diseases (2017ZZ02014), National Major Scientific and Technological Special Project for Significant New Drugs Development (2017ZX09304007), Key Projects in the National Science and Technology Pillar Program during the Thirteenth Five–year Plan Period (2018ZX09206005–004, 2017ZX10202202–005–004, 2017ZX10203201–008). All authors declared no competing interests. Anonymized datasets can be made available on reasonable request after approval from the trial management committee.

*Study registration:* ChiCTR2000029868

## INTRODUCTION

The coronavirus disease 2019 (COVID–19) caused by the severe acute respiratory syndrome coronavirus 2 (SARS–CoV–2) has swept into 185 countries/regions within five months. As of 22 April, more than 2.5 million infections and 178 thousand deaths have been reported.^1^

Several agents or drugs including, remdesivir, favipiravir, ribavirin, lopinavir–ritonavir (used in combination) and chloroquine (CQ) or hydroxychloroquine (HCQ), have been highlighted based on the promising *in–vitro* results and therapeutic experiences from another two coronavirus diseases including the severe acute respiratory syndrome and the Middle East respiratory syndrome.^2^ However, none of these promising results has yet been translated into clinical benefits of patients with COVID–19, including lopinavir–ritonavir, reported from the most recently failed trial.^3^

CQ and its hydroxy–analog HCQ, best known as antimalarial drugs, are glaring on the list of COVID–19 therapy, due to potent antiviral activity against SARS–CoV–2 from *in–vitro* studies,^4,5^ and promising results from news reports of some ongoing trials.^6^ Despite their unclear benefits, CQ and HCQ are both recommended for off–label use in the treatment of COVID–19 by the Chinese National guideline^7^ and recently authorized by the U.S. Food and Drug Administration for emergency use.^8^ HCQ was also recently recommended by the American president Donald Trump. Such a presidential endorsement stimulates an avalanche of demand for HCQ, which buried the dark–side of this drug. Deaths have been reported in Nigeria among people self–treating for apparent COVID–19 with CQ overdoses.^9^ Retinopathy, gastrointestinal, and cardiac side effects are well documented with the use of CQ or HCQ in the treatment of malarial and rheumatic diseases.^10^ HCQ is preferred in clinical applications due to its lower toxicity, particularly retinal toxicity,^10^ and three times the potency against SARS–CoV–2 infection comparing to CQ in the recent in–vitro study.^5^ Currently, there is no convincing evidence from well–designed clinical trials to support the use of CQ/HCQ with good efficacy and safety for the treatment of COVID–19. Rapidly conduction of such trials with high–quality is challenging in the face of a dangerous coronavirus outbreak, in which, healthcare workers are under overwhelming work and highest risk of exposure to developing COVID–19.^11^

Having encountered numerous challenges, we conducted a multicenter, open–label, randomized, controlled trial to assess the efficacy and safety of HCQ sulfate in adult patients with COVID–19. A clearer verdict will come from such a trial for the use of HCQ in patients with COVID–19.

## METHODS

### Trial oversight

The study was designed and initiated by the principal investigators after the protocol was approved by the institutional review board in Ruijin Hospital on February 6, 2020. The protocol and approval documents are available online with the full text of this article. Written informed consent was obtained from all patients. Hospitals with the capability of providing the current standard–of–care (SOC) for COVID–19 were invited to participate in the study by the principal investigators. Minimum requirements for the SOC included the provision of intravenous fluids, supplemental oxygen, regular laboratory testing, and SARS–CoV–2 test, hemodynamic monitoring, and intensive care, and the ability to deliver concomitant medications. The trial was conducted urgently during the outbreak of COVID–19 in compliance with the Declaration of Helsinki, Good Clinical Practice guidelines, and local regulatory requirements. The Shanghai Pharmaceuticals Holding Co.,Ltd donated the investigated drug, HCQ, but was not involved in the study design, accrual, analyses of data, or preparation of the manuscript. The principal investigators designed the study and made the decision to submit the manuscript for publication. All authors vouch for the trial protocol and fidelity of the study to the protocol, and veracity of the data and results. A contract research organization, R&G PharmaStudies Co., Ltd., was hired to assist in the study design and data collection, cleaning and statistical analyses. Data were recorded by clinical research coordinators followed by queries from clinical research associates. Confirmed data were then entered into the Web–based OpenClinica database for statistical analyses performed by the study statistician and reviewed by the senior statistician in accordance with Good Clinical Practice guidelines. Data collection forms and statistical analysis plans are available online with the full text of this article.

An independent data and safety monitoring committee (IDMC) periodically reviewed the progress and oversight of the study. The interim analysis was performed on March 14 and the results were presented to the IDMC. The rapid decline in eligible new cases of COVID–19 at that time in China precluded the recruitment of our targeted number of cases. After a two–round extensive review of the efficacy and safety data generated from the interim analysis, the IDMC endorsed an early termination of the trial. Members from the committee all agreed that the data from the trial is important for clinicians, the public, and the government to avoid inappropriate use of HCQ in the clinical management of COVID–19, particularly in overwhelming areas. The report of the trial could potentially be an important resource to facilitate a better design of future trials. The results of this clinical trial are reported in accordance with CONSORT (Consolidated Standards of Reporting Trials) guidelines.

### Trial design, Randomization, and procedures

This study was a multicenter, randomized, parallel, open–label, trial of HCQ in hospitalized patients with COVID–19. Patients were enrolled by the site investigators in 16 government–designated COVID–19 treatment centers from three provinces in China (Hubei, Henan and Anhui province). No placebo was used and drugs were not masked. Stratified random sampling was applied to stratify all eligible patients according to the disease severity (mild to moderate or severe) followed by random assignment (in a 1:1 ratio) in each stratum to ensure a balanced distribution of disease severity between treatment (HCQ plus SOC) and control (SOC only) groups. Randomization rules were designed by Leshan Liu together with principal investigators and implemented by an independent statistician who was not involved in data analysis. Equal numbers of cards with each group assignment number randomly generated by computer were placed in sequentially numbered envelopes that were opened as the patients were enrolled. All patients were managed with SOC aligning with the indications from the updating National clinical practice guidelines for COVID–19 in China. Patients in the treatment group were administrated with HCQ within 24 hours after randomization and were administrated with a loading dose of 1, 200 mg daily for three days followed by a maintained dose of 800 mg daily for remaining days (total treatment duration: 2 weeks or 3 weeks for mild/moderate or severe patients, respectively). The dose for HCQ was adjusted when adverse events were related to HCQ as judged by investigators. Details for dose adjustment were provided in the study protocol available online. Neither patients, nor investigators, nor statisticians were masked to treatment assignment. Lab technicians who performed virologic, chemistry, and other routine measurement were unaware of treatment information.

### Patients

Inclusion criteria: 1) age ≥18 years; 2) with ongoing SARS–CoV–2 infection confirmed in upper or lower respiratory tract specimens with real–time reverse–transcriptase–polymerase–chain–reaction (RT–PCR); 3) willingness to participate; 4) consent not to be enrolled by other clinical trials during the study period. Pneumonia on chest computed tomography was not mandatory for inclusion. Exclusion criteria: 1) age <18 years; 2) with severe conditions including malignancies, heart/liver/kidney disease or poorly controlled metabolic diseases; 3) not suitable to be administrated through the gastrointestinal tract;4) pregnant or lactation; 5) allergy to HCQ; 6) unable to cooperate with investigators due to cognitive impairments or poor mental status; 7) with severe liver disease (e.g. Child Pugh grade C, alanine aminotransferase >5 fold of times upper limit);8) with severe renal impairment (estimated glomerular filtration rate ≤30 mL/min/1.73 m^2^) or receiving continuous renal replacement therapy, hemodialysis, peritoneal dialysis. In the original protocol, patients with severe COVID–19 were excluded. Considering the anti–inflammatory property of HCQ might favor disease regression, we decided to include patients with severe COVID–19. (change approved by the Ethics Committee on February 17, 2020).

Definition for disease severity of COVID–19 was based on the 5^th^ version of Chinese guideline for the management of COVID–19^12^: Mild: patients with mild symptoms but no pneumonia manifestation on imaging; Moderate: patients with fever, cough, sputum production, other respiratory tract or non–specific symptoms along with pneumonia manifestation on imaging but no signs of severe pneumonia defined as the presence of SaO_2_/SPO_2_ <94% on room air or PaO_2_/FiO_2_ ratio ≤300mgHg.

### Assessment

Upper and/or lower respiratory tract specimens were obtained from each patient upon screening (Day –3~1), during treatment and post–treatment follow–up at scheduled visits on days 4, 7, 10, 14, 21 and 28. SARS–CoV–2 was measured by the local Center for Disease Control and Prevention or authorized health institutions or hospitals at each site using assays that have been approved by the National Medical Products Administration. Measurements were performed based on the recommendations by the National Institute for Viral Disease Control and Prevention (China) (http://ivdc.chinacdc.cn/kyjz/202001/t20200121_211337.html). Methods for total RNA extraction and amplification through RT–PCR were similar as described elsewhere.^13^ Instead of quantitative data (cycle threshold value) reported from RT–PCR assay, we only collected qualitative data reported from our trial sites. Based on national recommendation, a cycle threshold value less than 37 was defined as a positive test result, and a cycle threshold value of 40 or more was defined as a negative test. Cycle threshold values between 37 and 40 confirmed by retesting were reported as unclassified.

In addition to SARS–CoV–2, patients were assessed on each scheduled visit for vital signs, C–reactive protein, erythrocyte sedimentation rate, Tumor necrosis factor–α, Interleukin–6, complete blood cells count with differential, blood chemistry, coagulation panel, pulse oximetry, and respiratory symptoms. Administration records of HCQ and adverse events were reviewed daily to ensure fidelity to the protocol and more importantly, patient safety. Chest computed tomography was assessed upon screening and at the last visit of the treatment period (Day 14 for mild to moderate and Day 21 for severe patients). Chest computed tomography examinations can be exempted by the investigators if the participants can provide qualified examination results within 3 days before the initiation of the study. More details for data collection were provided in the protocol available online with the full text of this article.

### Outcome

The primary outcome for this trial was whether patients had a negative conversion of SARS–CoV–2 by 28 days and whether patients with severe COVID–19 had a clinical improvement by 28 days. However, since the trial was stopped early and only 2 patients with severe disease were enrolled, results on clinical improvement are not presented. We focused on the report of the primary outcome in this paper. Negative conversion of SARS–CoV–2 was defined as two consecutive confirmation of “Negatives” reported at least 24 hours apart without subsequent report of “Positive” SARS–CoV–2 by the end of the study. The date of the first “Negative” report was considered as the date of negative conversion. In the original protocol, the primary endpoint was prespecified as the “Negative conversion rate by Day10” (approved on February 6, 2020, by the Ethics Committee). However, with the increasing knowledge of COVID–19 from our clinical practice, we realized that the duration of SARS–CoV–2 in respiratory samples of many patients was longer than 10 days, recently highlighted by a detailed virologic study.^14^ We, therefore, modified our primary outcome to test whether patients had a negative conversion of SARS–CoV–2 by 28 days (approved on February 17, 2020, by the Ethics Committee). Meanwhile, negative conversion probability at Day4, 7, 10, 14, or 21 was specified as a secondary outcome as listed in the protocol but they do not appear on the trial registration list. The listed secondary outcome in the trial registration was adverse events coded using the latest version of Medical Dictionary for Regulatory Activities coding dictionary, recorded in standard medical terminology and graded according to the National Cancer Institute Common Terminology Criteria for Adverse Events.

Other pre–specified secondary outcomes not listed in the trial registration but included in the protocol were the probabilities of 1) alleviation of clinical symptoms; 2) improvement of C–reactive protein, erythrocyte sedimentation rate, tumor necrosis factor–α, Interleukin–6, and absolute blood lymphocyte count; 3) improvement of lung lesions on chest radiology; 4) all–cause death; 5) disease progression in mild to moderate patients. The time frame for these secondary outcomes were all from randomization to 28 days. Pre–specified secondary outcomes for severe patients were not listed here but in the protocol. Due to the early termination of our study, we could not able to justify the results from these analyses with an underpowered sample size and therefore decided not to emphasize them in this article to avoid misinterpretation. The only one presented here was the alleviation of clinical symptoms within 28 day which is an important outcome of interest prespecified in our protocol. Definition for the alleviation of clinical symptoms was 1) resolving from fever to an axillary temperature of ≤36.6 and; 2) normalization of SpO2 (>94% on room air) and; 3) disappearance of respiratory symptoms including nasal congestion, cough, sore throat, sputum production and shortness of breath.

### Statistical analysis

In the original protocol, the target number of enrollments was set to 200 without type–I error control. In the updated protocol (approved on February 10, 2020, by the Ethics Committee), we considered type–I and type–II error control and recalculated sample size. The sample size was calculated based on the alternative hypothesis of a 30% increase in the rate of conversion to negative of SARS–CoV–2 as defined by virus nucleic acid negativity. With the assumption that time to conversion follows an exponential distribution, the median time to conversion in HCQ can be reduced from 10 days to 7 days, therefore, a total of 248 events would provide a power of 80% to detect Hazards Ratio (HR)=0.7 (SOC vs. HCQ) with a Log–Rank test. Additionally, we assumed a 75–day accrual period and a 7–day follow–up following the last patient enrollment, therefore, approximately 360 subjects (180 per group) will be randomized in the study. An interim analysis was planned when around 150 patients were treated for at least 7 days. O’Brien–Fleming cumulative α–spending function by Lan–DeMets algorithm (Lan–Demets, 1983) was applied to control family–wise type–I error =0.05.

The overall negative conversion probability was estimated by analyzing time to negative conversion of SARS–CoV–2 using the Kaplan–Meier method in the intention–to–treat population and compared with a log–rank test. Patients failed to reach the negative conversion of SARS–CoV–2 (see definition in Outcome section) by the cutoff date of the analysis (14 March 2020) was considered as right–censored at the last visit date. All these patients remained in hospital and may reach negative conversion after our last visit date, but the specific timing of the event is unknown. Similar approaches were applied to the analysis of symptoms alleviation. Between–group rate difference was used to show treatment effect size and 95% confidence interval (CI) was estimated by an approximate normal distribution and the standard error was estimated by the bootstrap method (N=1000). The hazard ratio was estimated by the Cox model. HRs greater than 1 indicate the rate of virus negative conversion or symptoms alleviation is higher in the HCQ plus SOC group compared to the SOC group. Safety analyses were based on the patients’ actual treatment exposure. Data analyses were conducted on SAS version 9.4.

### Patient involvement

No patients were involved in setting the research question or the outcome measures, nor were they involved in developing plans for recruitment, design or implementation of the study. No patients were asked to advise on interpretation or writing up of results. There are no plans to disseminate the results of the research to study participants or the relevant patient community.

## RESULTS

### Patient

A total of 191 patients admitted with COVID–19 from February 11, 2020, to February 29, 2020, were assessed for eligibility, of which 41 did not meet eligibility criteria. The remaining 150 patients underwent randomization; Among them, 75 patients were assigned to SOC and 75 patients to SOC plus HCQ group (Figure 1). The mean age of the patients was 46 years and 82 (55%) were male. The mean days (± standard deviation, min to max) from symptoms onset to randomization was 16.6 (±10.5 days, 3 to 41 days) and 90 (60%) patients had concomitant medication before randomization; among them, 52 (34.7%) received antiviral treatment (Table 1). 148 (99%) patients had mild to moderate COVID–19 and only 2 (1%) patients were severe upon screening. Baseline demographic, epidemiological, and clinical characteristics of the patients between the two groups are shown in Table 1.

**Figure 1.**
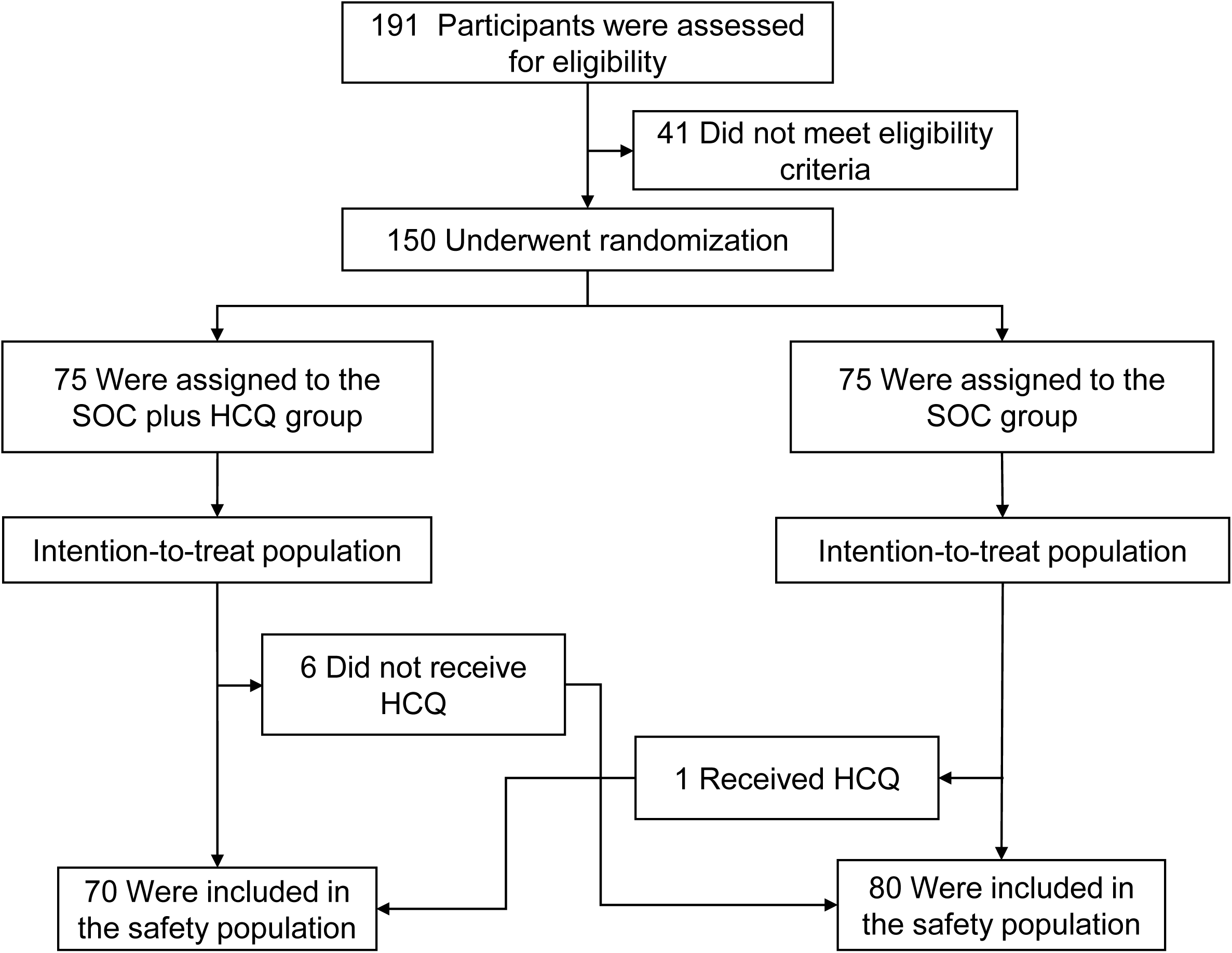
Screening, Randomization, and Follow–up. Shown is the disposition of the trial participants.

**Table 1.** Baseline Demographic and Clinical Characteristics of the Patients in the Intention–to–Treat Population.

| **Characteristics\*** | **SOC plus HCQ**<br>**(N=75)** | **SOC**<br>**(N=75)** | **Total**<br>**(N=150)** |
| --- | --- | --- | --- |
| Age — yr | 48.0±14.1 | 44.1±15.0 | 46.1±14.7 |
| Male sex — no. (%) | 42 (56.0) | 40 (53.3) | 82 (54.7) |
| Body mass index (% with missing data)<sup>†</sup> | 23.9±3.24 (1.3) | 23.2±3.0 (5.3) | 23.5±3.2 (3.3) |
| Days from disease onset to randomization (% with missing data) | 16.0±9.9 (2.7) | 17.1±11.1 (1.3) | 16.6±10.5 (2.0) |
| Exposure history — no./total no. (%) |  |  |  |
| Hubei province exposure | 50/72 (69.4) | 53/71 (74.6) | 103/143 (72) |
| Contact with confirmed COVID-19 patient (s) | 39/72 (54.2) | 32/71 (45.1) | 71/143 (49.7) |
| Others | 1/72 (1.4) | 1/71 (1.4) | 2/143 (1.4) |
| No exposure | 2/72 (2.8) | 9/71 (12.7) | 11/143 (7.7) |
| Unknown | 5/72 (6.9) | 5/71 (7) | 10/143 (7) |
| Medication prior to randomization — no. (%) | 47 (62.7) | 43 (57.3) | 90 (60.0) |
| Antivirals | 28 (37.3) | 24 (32.0) | 52 (34.7) |
| Arbidol | 12 (16.0) | 8 (10.7) | 20 (13.3) |
| Lopinavir-ritonavir | 18 (24.0) | 14 (18.7) | 32 (21.3) |
| Oseltamivir | 3 (4.0) | 3 (4.0) | 6 (4.0) |
| Entecavir | 1 (1.3) | 0 | 1 (0.7) |
| Virazole | 3 (4.0) | 6 (8.0) | 9 (6.0) |
| Ganciclovir | 0 | 2 (2.7) | 2 (1.3) |
| Disease severity — no. (%) |  |  |  |
| Mild | 15 (20.0) | 7 (9.3) | 22 (14.7) |
| Moderate | 59 (78.7) | 67 (89.3) | 126 (84.0) |
| Severe | 1 (1.3) | 1 (1.3) | 2 (1.3) |
| Coexisting conditions — no./total no. (%) | 28 (37.3) | 17 (22.7) | 45 (30.0) |
| Diabetes | 12 (16.0) | 9 (12.0) | 21 (14.0) |
| Hypertension | 6 (8.0) | 3 (4.0) | 9 (6.0) |
| Others | 21 (28.0) | 10 (13.3) | 31 (20.7) |
| Vital Signs (% with missing data) |  |  |  |
| Body temperature — °C | 36.9±0.47 (4) | 36.8±0.48 (0.0) | 36.8±0.5 (2.0) |
| Pulse — beats/min | 82.75±8.0 (2.7) | 82.5±9.4 (5.3) | 82.6±8.7 (4.0) |
| Respiratory rate — breaths/min | 19.6±1.3 (2.7) | 19.7±1.7 (6.7) | 19.6±1.5 (4.7) |
| Systolic blood pressure — mm Hg | 126.3±13.2 (6.7) | 123.5±11.2 (8.0) | 124.9±12.3 (7.3) |
| Diastolic blood pressure — mm Hg | 79.1±8.5 (6.7) | 76.8±8.0 (8.0) | 77.9±8.3 (7.3) |
| Pulse oximetry — % | 97.4±1.6 (0) | 97.3±1.6 (2.7) | 97.4±1.6 (1.3) |
| Symptoms — no./total no. (%) |  |  |  |
| Fever | 43/72 (59.7) | 40/75 (53.3) | 83/157 (52.9) |
| Cough | 35/68 (51.5) | 26/68 (38.2) | 61/136 (44.9) |
| Sputum production | 11/68 (16.2) | 4/68 (5.9) | 15/136 (11) |
| Shortness of breath | 15/68 (22.1) | 4/68 (5.9) | 19/136 (14) |
| Nasal congestion | 0 (0) | 0 (0) | 0 (0) |
| Pharynx discomfort | 2/68 (2.9) | 4/68 (5.9) | 6/136 (4.4) |
| Fatigue | 5/68 (7.4) | 1/68 (1.5) | 6/136 (4.4) |
| Laboratory parameters (% with missing data) |  |  |  |
| White cell count — $\times 10^9$/liter | 5.59±1.9 (0) | 5.6±1.8 (0.0) | 5.6±1.8 (0.0) |
| Lymphocyte count — $\times 10^9$/liter | 1.46±0.6 (0) | 1.6±0.5 (0.0) | 1.5±0.57 (0.0) |
| Neutrophil count — $\times 10^9$/liter | 3.55±1.6 (0) | 4.2±6.2 (0.0) | 3.9±4.51 (0.0) |
| Platelet count — $\times 10^9$/liter | 214.8±68.1 (0) | 211.7±71.6 (0.0) | 213.2±69.7 (0.0) |
| Hemoglobin — g/liter | 128.8±17.5 (0) | 129.1±17.1 (0.0) | 129.0±17.3 (0.0) |
| Aspartate aminotransferase — U/liter | 25.0±13.5 (0) | 26±14.7 (0.0) | 25.5±14.1 (0.0) |
| Alanine aminotransferase — U/liter | 31.4±26.3 (0) | 32.7±25.2 (1.3) | 32.1±25.7 (0.7) |
| γ-glutamyl transpeptidase — U/liter | 46.9±61.8 (2.7) | 44.0±51.8 (2.7) | 45.4±56.9 (2.7) |
| Total bilirubin — μmol/liter | 11.6±8.4 (1.3) | 12.8±7.7 (2.7) | 12.2±8.1 (2.0) |
| Albumin — g/L | 39.9±4.5 (1.3) | 40.4±4.4 (1.3) | 40.1±4.4 (1.3) |
| Lactate dehydrogenase — U/liter | 203.9±65.2 (12) | 190.9±49.5 (10.7) | 197.4±58.0 (11.3) |
| Creatine kinase — U/liter | 74.4±110.1 (10.7) | 71.0±52.6 (9.3) | 72.7±85.7 (10.0) |
| Creatine kinase isoenzyme-MB — U/liter | 8.0±4.2 (38.7) | 6.8±3.9 (41.3) | 7.4±4.0 (40.0) |
| Creatinine — μmol/liter | 71.2±38.4 (1.3) | 63.9±16.0 (1.3) | 67.5±29.5 (1.3) |
| Blood urea nitrogen — mmol/liter | 3.5±1.0 (41.3) | 3.1±0.7 (48.0) | 3.3±0.9 (44.7) |
| Urea — mmol/liter | 4.0±3.0 (58.7) | 3.8±1.2 (57.3) | 4.0±2.2 (58.0) |
| International normalized ratio | 1.0±0.1 (2.7) | 1.0±0.1 (1.3) | 1.0±0.1 (2.0) |
| C-reactive protein mg/liter | 9.9±13.3 (2.7) | 7.4±12.8 (1.3) | 8.6±13.1 (2.0) |
| Erythrocyte sedimentation rate | 30.6±28.6 (4) | 25.4±21.7 (5.3) | 28.0±25.4 (4.7) |
| TNF-α — pg/milliliter | 4.9±4.1 (90.7) | 4.8±3.6 (90.7) | 4.8±3.7 (90.7) |
| IL-6 — pg/milliliter | 12.9±36.3 (58.7) | 8.9±13.0 (61.3) | 11.0±27.4 (60.0) |

\* Plus-minus values are means ± SD. SOC=standard-of-care, HCQ= hydroxychloroquine, COVID-19=coronavirus disease 2019. To convert the values for creatinine to milligrams per deciliter, divide by 88.4. To convert the values for total bilirubin to milligrams per deciliter, divide by 17.1.
† The body mass index is the weight in kilograms divided by the square of the height in meters.

By 14 March 2020 (the cutoff date for data analysis), the median duration of follow–up was 21 days (range, 2 to 33 days) in the SOC group and 20 days (range, 3 to 31 days) in the SOC plus HCQ group with 9 patients in each group having complete 28-day follow-up. Of the 75 patients assigned to receive SOC plus HCQ, 6 patients did not receive any dose of HCQ; of them, 3 patients withdrew consent and 3 patients refuse to be administrated HCQ. The co–intervention including antiviral, antibiotics, and systemic glucocorticoid therapy between the two groups were similar (Table 2). One patient with the moderate disease in the HCQ group progressed to severe COVID–19 and no patients died during follow–up.

**Table 2.** Treatments after Randomization in Patients in the Intention–to–Treat Population.

| **Treatments** | SOC plus HCQ | SOC | Total |
| --- | --- | --- | --- |
|  | (N=75) | (N=75) | (N=150) |
| Antiviral — no. (%) | 47 (62.7) | 48 (64.0) | 95 (63.3) |
| Arbidol | 37 (49.3) | 33 (44.0) | 70 (46.7) |
| Virazole | 13 (17.3) | 15 (20.0) | 28 (18.7) |
| Lopinavir–Ritonavir | 13 (17.3) | 12 (16.0) | 25 (16.7) |
| Oseltamivir | 8 (10.7) | 9 (12.0) | 17 (11.3) |
| Entecavir | 1 (1.3) | 1 (1.3) | 2 (1.3) |
| Antibiotics — no. (%) | 32 (42.7) | 27 (36.0) | 59 (39.3) |
| Systemic glucocorticoid therapy — no. (%) | 6 (8.0) | 4 (5.3) | 10 (6.7) |

SOC=standard-of-care, HCQ= hydroxychloroquine

### Primary Outcome

A total of 109 (72.7%) patients (56 in SOC and 53 in SOC plus HCQ) negatively converted well before 28 days and the remaining 41 (27.8%) patients (19 in SOC and 22 in SOC plus HCQ) were censored due to the failure of reaching virus negative conversion. The maximum duration for a patient with positive SARS–CoV–2 was 23 days by the cutoff date of our analysis. Overall, the negative conversion probability of SARS–CoV–2 among patients who were assigned to receive SOC plus HCQ was 85.4% (95%CI 73.8% to 93.8%), similar to that of the SOC group 81.3% (95%CI 71.2% to 89.6%) by 28 days. The difference of negative conversion probability between SOC plus HCQ and SOC alone was 4.1% (95%CI –10.3% to 18.5%). The median time to negative conversion was also similar in the SOC plus HCQ group (8 days, 95%CI 5 to 10 days) with that in the SOC group (7 days, 95%CI 5 to 8 days) (Hazard ratio, 0.846; 95%CI, 0.58 to 1.23; p=0.34 by log–rank test) (Figure 2).

**Figure 2.**
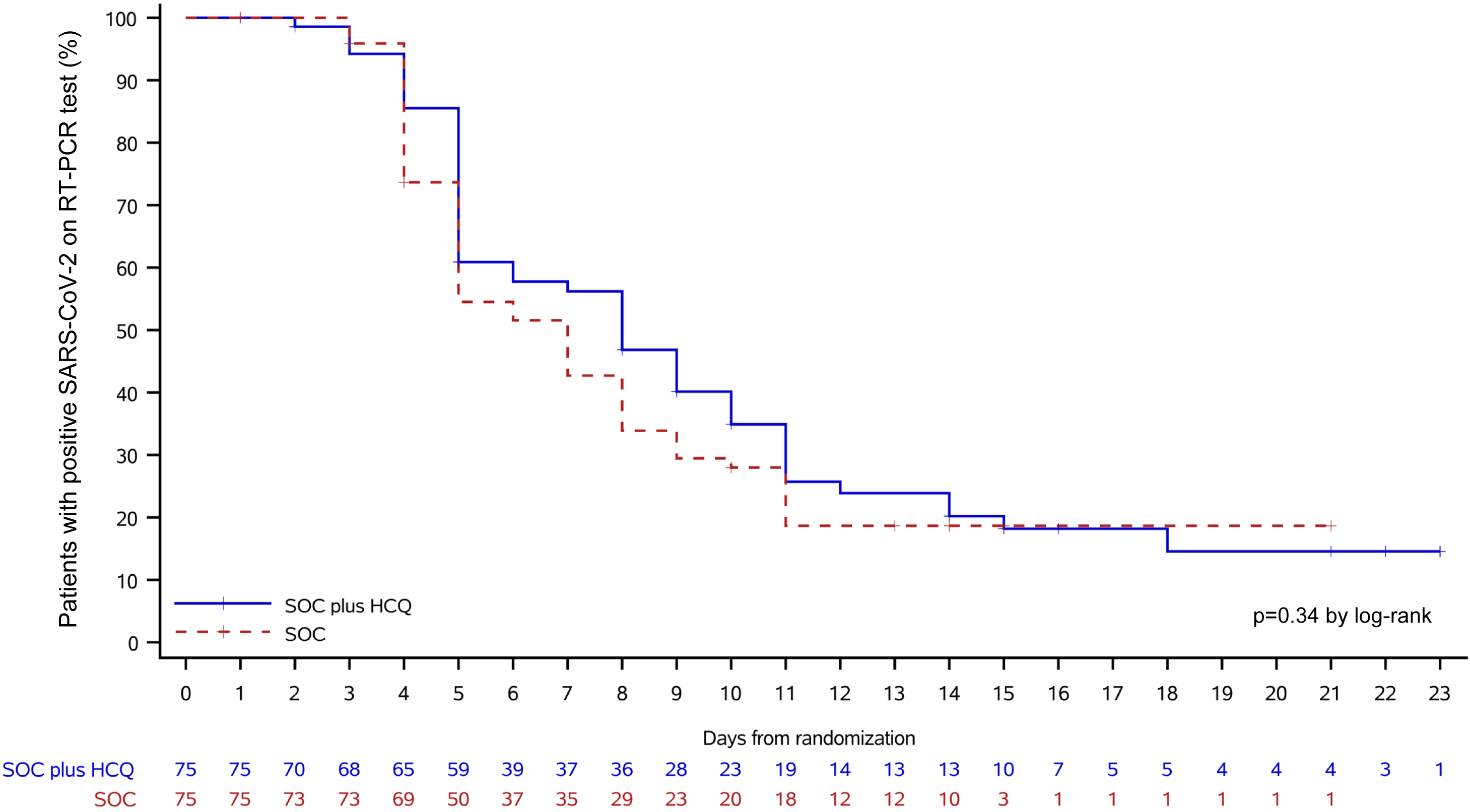
Kaplan–Meier curves of the time to negative conversion of SARS–CoV–2 on RT–PCR test in SOC plus HCQ *versus* SOC in the intention–to–treat population. Shown are data for 75 patients assigned to SOC plus HCQ and 75 assigned to SOC. The overall negative conversion probability was 85.4% (95% CI 73.8% to 93.8%), similar to that of the SOC group 81.3% (95%CI 71.2% to 89.6%) by 28 days (p=0.34). Between-group difference was 4.1% (95% CI –10.3% to 18.5%). The median time to negative conversion was also similar in the SOC plus HCQ group (8 days, 95% CI 5 to 10 days) with that in the SOC group (7 days, 95% CI 5 to 8 days) (Hazard ratio, 0.846; 95% CI, 0.58 to 1.23; p=0.34 by log-rank test). Data from patients who did not have negative conversion were censored (tick marks) at the last visit date. RT–PCR=real time reverse transcription polymerase chain reaction; SOC=standard–of–care; HCQ=hydroxychloroquine; CI=confidence interval.

### Safety

Six patients assigned to the SOC plus HCQ group but did not receive HCQ treatment were classified as HCQ non–recipient in the safety population. One patient in the SOC group wrongly received 14–day of HCQ treatment with an accumulative dose of 11, 600 mg. This patient was classified as HCQ recipient in the safety population (Figure 1). Safety endpoints were compared between HCQ recipient and non–recipient (Table 3). In HCQ recipients, the median duration of HCQ treatment was 14 days (range, 1 to 22). Between randomization and final visit, a total of 21 (30%) patients in the SOC plus HCQ group reported adverse events, higher than those 7 (8.8%) patients reported in the SOC group (Table 3). No serious adverse events were reported in the SOC group. Two patients in the HCQ group reported serious adverse events due to disease progression and upper respiratory infection. The case with upper respiratory infection discharged after finishing the 14–day treatment of HCQ and developed throat–drying and pharyngalgia requiring re–admission without evidence of pneumonia on chest computed tomography during the extended follow–up period.

**Table 3.** Summary of Adverse Events in the Safety Population.

| Adverse Events* | SOC plus HCQ | SOC |
| --- | --- | --- |
|  | (N=70) | (N=80) |
|  | no. of patients (%) |  |
| Any adverse event | 21 (30) | 7 (8.8) |
| Serious adverse event | 2 (2.9) | 0 |
| Disease progression | 1 (1.4) | 0 |
| Upper respiratory tract infection | 1 (1.4) | 0 |
| Non serious adverse event | 19 (27.1) | 7 (8.8) |
| Diarrhea | 7 (10.0) | 0 |
| Vomiting | 2 (2.9) | 0 |
| Nausea | 1 (1.4) | 0 |
| Abdominal discomfort | 1 (1.4) | 0 |
| Thirst | 1 (1.4) | 0 |
| Abdominal bloating | 0 | 1 (1.3) |
| Sinus bradycardia | 1 (1.4) | 0 |
| Hypertension | 1 (1.4) | 0 |
| Orthostatic hypotension | 1 (1.4) | 0 |
| Hypertriglyceridemia | 1 (1.4) | 0 |
| Decreased appetite | 1 (1.4) | 0 |
| Fatigue | 1 (1.4) | 0 |
| Fever | 0 | 1 (1.3) |
| Dyspnea | 1 (1.4) | 0 |
| Flush | 1 (1.4) | 0 |
| Liver abnormality | 0 | 1 (1.3) |
| Kidney injury | 1 (1.4) | 0 |
| Coagulation dysfunction | 1 (1.4) | 0 |
| Hepatic steatosis | 0 | 1 (1.3) |
| Otitis externa | 0 | 1 (1.3) |
| Blurred vision | 1 (1.4) | 0 |
| Decreased white blood cell | 1 (1.4) | 0 |
| Increased alanine aminotransferase | 1 (1.4) | 1 (1.3) |
| Increased serum amylase | 1 (1.4) | 0 |
| Decreased neutrophil count | 1 (1.4) | 0 |
| Increased serum amyloid A | 0 | 1 (1.3) |

\* Multiple occurrences of the same adverse event in one patient were counted. SOC=standard-of-care, HCQ=hydroxychloroquine.

The most common adverse events in the SOC plus HCQ group were diarrhea reported in 7 (10%) patients, which was not reported in the SOC group. HCQ was discontinued in one patient due to blurred vision and was adjusted to give a lower dose in one patient who reported thirst. These two adverse events were both transient with a period of 1–2 days.

### Secondary Outcome

Negative conversion probability by a specific time–point, 4–, 7–, 10–, 14– or 21–day was also similar between the two groups (Supplementary Table 1). The probability of symptoms alleviation by 28 days was similar between patients with SOC with (59.9%, 95%CI 45.0% to 75.3%) and without HCQ (66.6%, 95%CI 39.5% to 90.9%). Between–group difference was –6.6% (95%CI–41.3% to 28.0%). The median time to alleviation of clinical symptoms was similar in the SOC plus HCQ group with that in the SOC group (19 days *versus* 21 days, Hazard ratio, 1.01, 95%CI, 0.59 to 1.74, p=0.97 by log–rank test) (Figure 3).

**Figure 3.**
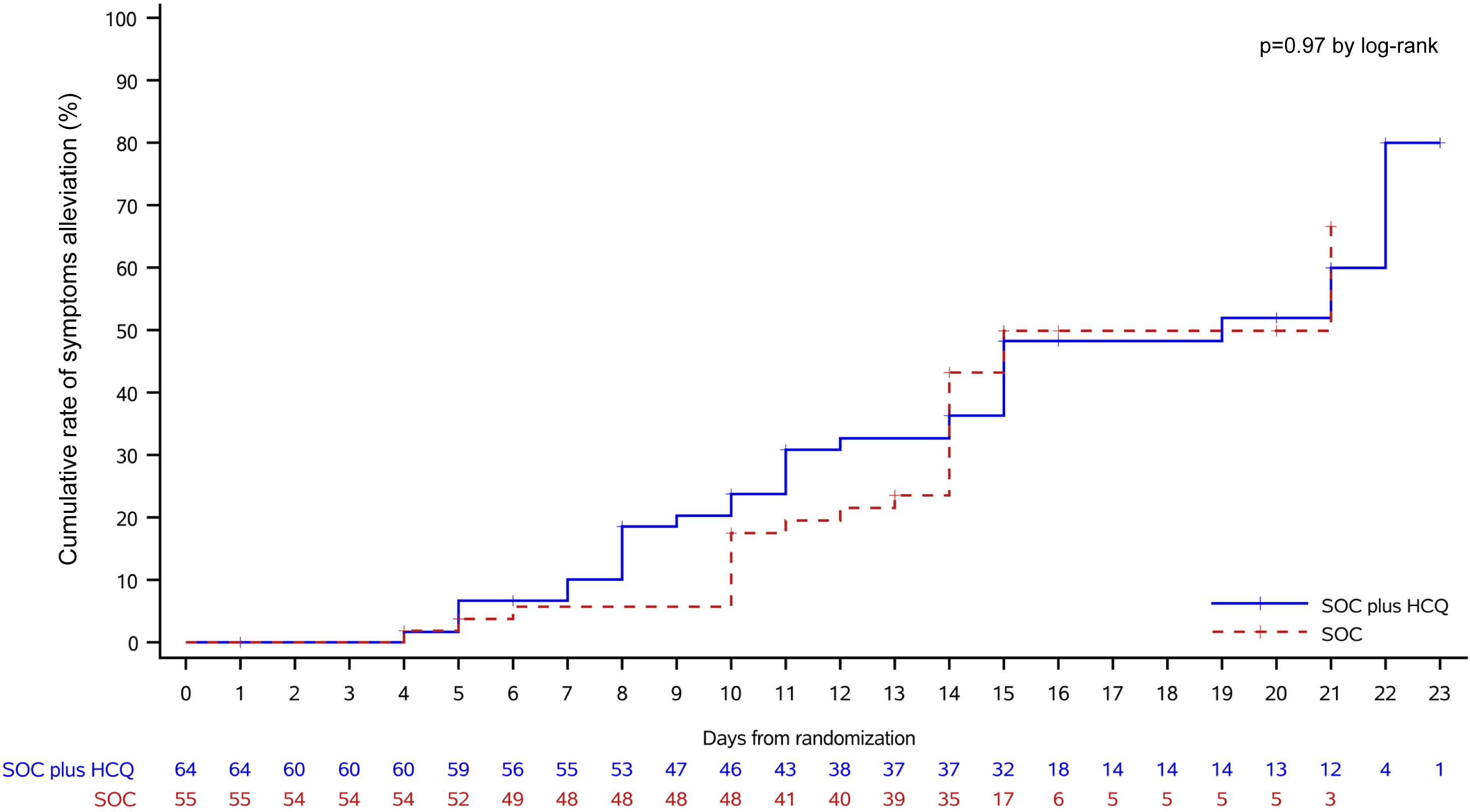
Kaplan–Meier curves of the time to alleviation of clinical symptoms in SOC plus HCQ *versus* SOC in the intention–to–treat population. Shown are data for 55 symptomatic patients assigned to SOC plus HCQ and 64 symptomatic patients assigned to SOC. The probability of symptoms alleviation by 28 days was similar (p=0.97) between patients with SOC with (59.9%, 95%CI 45.0% to 75.3%) and without HCQ (66.6%, 95%CI 39.5% to 90.9%). Between-group difference was –6.6% (95%CI –41.3% to 28.0%). The median time to alleviation of clinical symptoms was similar in the SOC plus HCQ group with that in the SOC group (19 days *versus* 21 days, Hazard ratio, 1.01, 95%CI, 0.59 to 1.74, p=0.97 by log-rank test). Data from patients who did not have symptoms alleviation were censored (tick marks) at the last visit date. SOC=standard–of–care; HCQ=hydroxychloroquine; CI=confidence interval.

## DISCUSSION

### Principal findings

The present study (conducted during the outbreak of COVID–19 in China) is the first randomized, controlled trial evaluating HCQ administration in COVID–19 patients. The findings do not provide evidence to support an increase of negative conversion probability of SARS–CoV–2 conferred by the addition of HCQ administration to the current SOC in patients hospitalized mainly with persistent mild to moderate COVID–19.

### Comparison with other studies

Our negative results on the antiviral efficacy of HCQ obtained in this trial are on the contrary to the encouraging *in–vitro* results^4,5^ and the recently reported promising results from a non–randomized trial with 36 COVID–19 patients.^6^ It should be noted that participants in our trial had mainly mild to moderate disease with a median 16–day delay of HCQ treatment from symptoms onset. Therefore, the negative results of our trial are only applicable to patients with persistently mild to moderate COVID–19. The COVID–19 has overwhelmed hospital systems during the pandemic, many of these patients may be treated in the community and may finally end up in hospitalization because of oxygen needs or even rapid deterioration of the disease. Current data from our trial do not provide evidence to support the use of HCQ in this particular population, particularly considering the increased adverse events (discussed below). Our trial could not answer the antiviral efficacy of HCQ at the earlier stage, e.g., within 48h of illness onset, the golden window for antiviral treatment in influenza.^16^ However, it is challenging to conduct such a trial in hospitalized patients and will be easier in outpatient or community settings. The fact that no restriction of antiviral treatment in our trial should also be considered when interpreting our results. It would be more conclusive to see the antiviral effects of HCQ when comparing a “pure” control arm. But in the early dangerous outbreak of COVID–19 in China, it is not quite ethical to set a restriction on drugs that could be potentially useful. Nevertheless, the usage of antiviral before and after randomization between the HCQ and SOC groups was balanced and therefore might affect little on our primary endpoint. Moreover, it is not likely to have additional antiviral effects by further escalating the dosage of HCQ used in our trial, because that the dosage of HCQ we choose can reach the 50% effective concentrations (EC_50_) of HCQ against SARS–CoV–2,^16^ although we did not monitor the concentration of HCQ in our study. Another important message from our trial is that the K–M curve crossed over with time, suggesting a potential for a non–constant hazard ratio of negative conversion conferred by the exposure to HCQ. Therefore, the HRs presented in our trial should be interpreted as weighted results over time rather than a definitive constant. We will investigate it in the future. Further trials might also need to combat this issue when using the Cox regression method. Taken together, future studies could take advantage of our results to design trials in more selective populations, at the earliest stage as possible (<48h of illness onset), and using more sensitive endpoints, such as viral load shedding.

HCQ in our trial was given with a loading dose of 1, 200 mg daily for three days followed by a maintained dose of 800 mg daily for a total treatment duration of 2 weeks or 3 weeks for mild/moderate or severe patients, respectively. Serious adverse events occurred in 2 patients and both were reported from HCQ recipients. The overall frequency of adverse events was significantly higher in HCQ recipients than non–recipients. Gastrointestinal events, particularly diarrhea, was most commonly reported, similar to another report using a high dose of HCQ^19^ Transient blurred vision was reported in one patient whose symptoms recovered 2 days after discontinuation of HCQ. Early development of retinal damage with a daily dose of 800 to 1,200mg was detected using sensitive retinal screening tests.^20^ Therefore, the retinal damage could be underestimated in our trial. Events of cardiac arrhythmia, *e.g*., prolonged QT interval^21^ was not observed in our trial, possibly due to the relatively mild/moderate patients investigated or the short–term period of follow–up. However, with the increasing interest of the combined use of HCQ and azithromycin worldwide, physicians should be cautious of the increased risk of QT interval prolongation and fatal ventricular arrhythmia with azithromycin and other antimicrobials.^22,23^ Drug–drug interaction^10^ should be taken into consideration when assessing safety and efficacy endpoints in future HCQ trials. The effects of HCQ in causing increased levels of digitoxin and metoprolol^17^ would be particularly relevant in severe COVID–19 patients and therefore would require close monitoring.

### Strengths and limitations of this study

This study provides the first and timely evidence regarding benefit–risk of HCQ derived from a multi–center randomized controlled trial, which was initiated during the most challenging time of the COVID–19 outbreak in China.

Under such a situation, our study does have several limitations. First, the design of open–label, as opposed to double–blind design, introduces biased investigator–determined assessments and unbalanced dosage adjustment. Urgent production of placebos mimicking HCQ and the management of a multi–center placebo–controlled trial remains challenging during the pandemic. Second, the use of sequential envelopes is inferior to the interactive web response management system for randomization. Third, the conduction of our trial in the setting of hospitalized patients precludes us from enrolling patients at the early disease stage. In addition, we cannot provide evidence on the utility of HCQ regarding disease progression or regression because 148 out of 150 (99%) patients in our trial are with mild to moderate disease. Fourth, the results on our main pre–specified outcomes are not entirely conclusive based on underpowered sample size due to the lack of enough eligible patients to enroll. The recruitment of eligible patients was unexpectedly difficult with almost hundreds of clinical trials launched in the same period in response to the urgent call for the exploration of effective treatment against COVID–19 by the national health authorities. The rapid decline in eligible new cases owing to the successful containment of COVID–19 in the mid of March 2020 in China precluded further recruitment to reach our targeted sample size. The premature termination of our trial also led to increased censoring data on our primary outcome. The probability of negative conversion between the two groups was therefore re–analyzed using follow–up data by April 27, 2020 (Supplementary Table 2) and the results were consistent with our current results. Fifth, it is difficult to ensure the fidelity to the protocol by site investigators under highly challenging circumstances at the front lines in the COVID–19 treatment centers. Hiring a contract research organization, as we did in our trial, can greatly support the conduct and oversight of the trial. Sixth, population quarantine of Wuhan and neighboring cities, nationwide travel restrictions, and case/contact isolation were also barriers to collect and transfer data and paper files. Several prespecified secondary endpoints, including the imaging changes on chest CT, were therefore not finished by the cutoff date of analysis. Finally, the specimens collected in our trial for virus RNA determination are mostly from the upper respiratory tract rather than bronchoalveolar lavage fluid, which introduced false–negative results.^24^ But the prespecified definition for virus negative conversion was two consecutive negatives with at least 24 hours apart, which can reduce false negativity.

### Conclusion and policy implications

The results of our trial did not show additional benefits of virus elimination by adding HCQ to the current SOC in patients mainly with persistent mild to moderate COVID–19. Adverse events, particularly gastrointestinal events, were more frequently reported in patients receiving HCQ, who were given with a loading dose of 1, 200 mg daily for three days followed by a maintained dose of 800 mg daily for remaining days (total treatment duration: 2 weeks or 3 weeks for mild/moderate or severe patients, respectively). Overall, these data do not support adding HCQ to the current SOC in patients with persistent mild to moderate COVID–19 for eliminating the virus. Our trial may provide initial evidence for the benefit–risk of HCQ and serve as a resource to support further research.

## Data Availability

Anonymized datasets can be made available on reasonable request after approval from the trial management committee and after signing a data access agreement. Proposals should be directed to the corresponding author.

## Acknowledgments

We would like to acknowledge the members of the trial steering committee for their many contributions in conducting the trial under very challenging field conditions, in particular, Prof. Yiping Xu and the members of the independent data and safety monitoring committee (Prof. Shenghan Lai [chair], Prof. Weimin Li, Prof. Hejian Zou, Prof. Xinxin Zhang, and Prof. Gang Chen) for their oversight and the Chief of the Department of Anesthesiology, Ruijin Hospital, Shanghai Jiao Tong University School of Medicine, Prof. Yan Luo for her support to the trial. We would like to acknowledge the contribution of Xia Chen, Li Li and Jian Li for their support of statistical analysis. We also would like to acknowledge the contribution of the other members of the coordinating centers: Ming Li, Yongcai Gao, Ming Wu, Zuohan Xiao, Shanshan Deng, Xu Zhang, Xiaobo Chen, Haiyan Hang, Haiguang Xin, Yadong Gao, Dandan Dong, Mingjie, Hou, Qiuping Peng, Yiming Yan, Peng Gao, Lin Sun, Jingdong Shi, Hong Tang, and Fei Deng. Most importantly, we thank the donation of our investigated drug from the Shanghai Pharmaceuticals Holding Co., Ltd and the patients themselves for their bravery and altruism in participating in this trial.

## Funding

This work was supported by the Emergent Projects of National Science and Technology (2020YFC0844500), National Natural Science Foundation of China (81970020, 81770025), National Key Research and Development Program of China (2016YFC0901104), Shanghai Municipal Key Clinical Specialty (shslczdzk02202, shslczdzk01103), National Innovative Research Team of High–level Local Universities in Shanghai, Shanghai Key Discipline for Respiratory Diseases (2017ZZ02014), National Major Scientific and Technological Special Project for Significant New Drugs Development (2017ZX09304007), Key Projects in the National Science and Technology Pillar Program during the Thirteenth Five–year Plan Period (2018ZX09206005–004, 2017ZX10202202–005–004, 2017ZX10203201–008). The funders played no role in study design, data collection, data analysis, data interpretation, or reporting. The guarantors had full access to all the data in the study, take responsibility for the integrity of the data and the accuracy of the data analysis, and had final responsibility for the decision to submit for publication.

## Competing interests

All authors have completed the ICMJE uniform disclosure form at www.icmje.org/coi_disclosure.pdf and declare: no support from any organization for the submitted work; no financial relationships with any organizations that might have an interest in the submitted work in the previous three years; no other relationships or activities that could appear to have influenced the submitted work.

## Ethical approval

The trial was approved by the Institutional Review Board of Ruijin Hospital, Shanghai Jiao Tong University School of Medicine (KY2020–29).

## Patient consent

Written informed consent was obtained.

## Transparency declaration

The lead author (the manuscript’s guarantor) affirms that the manuscript is an honest, accurate, and transparent account of the study being reported; that no important aspects of the study have been omitted; and that any discrepancies from the study as planned (and, if relevant, registered) have been explained.

## Dissemination to participants and related patient and public communities

This is an Open Access article distributed in accordance with the Creative Commons Attribution Non Commercial (CC BY-NC 4.0) license, which permits others to distribute, remix, adapt, build upon this work non-commercially, and license their derivative works on different terms, provided the original work is properly cited and the use is non-commercial. See: http://creativecommons.org/licenses/by-nc/4.0/.

This manuscript been deposited at medrxiv as a preprint (DOI: https://doi.org/10.1101/2020.04.10.20060558)

## Notes

### Competing Interest Statement

The authors have declared no competing interest.

### Clinical Trial

ChiCTR2000029868

